# The Silent Pandemic COVID-19 in the Asymptomatic Population

**DOI:** 10.1101/2020.12.29.20248985

**Authors:** Robert L. Stout, Steven J. Rigatti

## Abstract

As the COVID-19 pandemic continues to ravage the world there is a great need to understand the dynamics of spread. Currently the seroprevalence of asymptomatic COVID-19 doubles every 3 months, this silent epidemic of new infections may be the main driving force behind the rapid increase in SARS-CoV-2 cases.

Public health official quickly recognized that clinical cases were just the tip of the iceberg. In fact a great deal of the spread was being driven by the asymptomatically infected who continued to go out, socialize and go to work. While seropositivity is an insensitive marker for acute infection it does tell us about the prevalence COVID-19 in the population.

**Objective:** Describe the seroprevalence of SARS-CoV-2 infection in the United States over time.

**Methodology:** Repeated convenience samples from a commercial laboratory dedicated to the assessment of life insurance applicants were tested for the presence of antibodies to SARS-CoV-2, in several time periods between May and December of 2020. US census data were used to estimate the population prevalence of seropositivity.

**Results:** The raw seroprevalence in the May-June, September, and December timeframes were 3.0%, 6.6% and 10.4%, respectively. Higher rates were noted in younger vs. older age groups. Total estimated seroprevalence in the US is estimated at 25.7 million cases.

**Conclusions:** The seroprevalence of SARS-CoV-2 demonstrates a significantly larger pool of individuals who have contract COVID-19 and recovered, implying a lower case rate of hospitalizations and deaths than have been reported so far.

## Manuscript

SARS-CoV-2 was first recognized in the US in January^1^. Since then, as of December 6^th^, 2020, 14.8 million additional cases have been reported to Center for Disease Control and Prevention^2^. Because a substantial proportion of COVID-19 infections may be asymptomatic or only mildly symptomatic, these reported cases may substantially underestimate the true cumulative prevalence of infection. Except for severe cases the symptoms for SARS-CoV-2 are easily confused with head colds, allergy, and the flu^3^. In addition, symptomatic patients many have chosen to avoid testing or had no access to testing, both resulting in an underestimation of the severity of the pandemic.

Antibodies against SARS-CoV-2 have been noted to persist for months, and thus studies of seroprevalence may provide a more reliable estimate of the dimensions of the current pandemic.

## Methods

In the United States, the process of purchasing life insurance often involves testing of blood and urine specimens for common tests related to overall health. Individuals applying for life insurance are a self-selected group primarily from middle and higher socio-economic strata. Those who have a history of chronic illness may be less likely to apply because more serious conditions can be associated with higher life premiums.

In three time periods between May12 and December 6 2020, a total of 126,587 individuals were tested for antibodies to SARS-CoV-2. Individuals were part of a convenience sample from a pool of life insurance applicants who had blood tests performed as part of life insurance underwriting at Clinical Reference Laboratories. This sample represents approximately one fifth of all samples tested at the facility during those time frames. All applicants self-reported that they were well at the time of application. The antibody tests were performed using the Roche Elecsys Anti-SARS-CoV-2 kit on the Roche 602 analyzer, with a stated sensitivity of 100% and specificity of 99.8%, utilizing an electrochemiluminescence immunoassay.

The differences in continuous variables between the antibody-positive and negative groups were tested for significance with the Mann-Whitney U test, while differences in categorical variables were tested using the chi-square test.

To estimate the total burden of SARS-CoV-2 infections in the US, census data was obtained. For each state and the District of Columbia, the total 2019 estimated census population was multiplied by the US population proportion between the ages of 16 and 84 (77%). Then, the state-specific proportion of positive tests was applied from our sample. Confidence limits were estimated by generating 5000 bootstrap samples (with replacement) of our data and recalculating the total number of US cases. Under and over-representation of states was determined by a ratio between the proportion of individuals living in a given state to the proportion of tests performed in that state.

All statistical analyses were performed using R (version 3.6.1)^4^ and R-studio (version 1.2.1335)^5^. This study design was approved by the Western Institutional Review Board and determined to be exempt under the Common Rule and applicable guidance and determined it is exempt under 45 CFR § 46.104(d)(4) using de-identified study samples for epidemiologic investigation, WIRB Work Order #1-1324846-1.

## Results

The overall rate of SARS-CoV-2 antibody positivity was 5.64%. The seronegative group was noted to be significantly older and to have a higher proportion of males (Table 1). Rates of seropositivity trended downward across age, and upward over time. The rates among the youngest group were 3.6% in May/June, 9.8% in September and 15.5% in December, with lower rates of increase in older age groups (see Table 2 and Figure 1). This alarming trend seems to coincide with higher rates of asymptomatic infection and less stringent practice of social distancing in the younger age group.

**Table 1:** Characteristics of study population by SARS-CoV-2 antibody status.

|  | **SARS-CoV-2 negative**<br>n = 120763 | **SARS-CoV-2 positive**<br>n = 7232 | **p** |
| --- | --- | --- | --- |
| Age (yrs), median[IQR] | 44 [34,54] | 41 [32,50] | <0.001<sup>1</sup> |
| 16-40 | 45.30% | 51.90% |  |
| 41-60 | 41.40% | 40.00% |  |
| 61-85 | 13.30% | 8.00% |  |
| Sex, (% male) | 55.80% | 54.20% | 0.0021 |

<sup>1</sup>p-value by Pearson Chi-square test

**Table 2:** SARS-CoV-2 Seropositivity Rate

| **Age** | **May/June** | **September** | **December** | **Total** |
| --- | --- | --- | --- | --- |
| <30 | 3.6% | 9.8% | 15.5% | 7.9% |
| 30 - 39 | 3.1% | 6.7% | 10.9% | 5.8% |
| 40 - 49 | 3.2% | 6.7% | 10.2% | 5.8% |
| 50 - 59 | 2.9% | 5.9% | 9.2% | 5.1% |
| 60 - 69 | 2.6% | 4.4% | 7.7% | 4.0% |
| ≥70 | 1.4% | 2.8% | 4.1% | 2.4% |
| Total | 3.0% | 6.6% | 10.4% | 5.6% |
| Tested | 50,072 | 61,910 | 14,605 | 126,587 |

**Figure 1:**
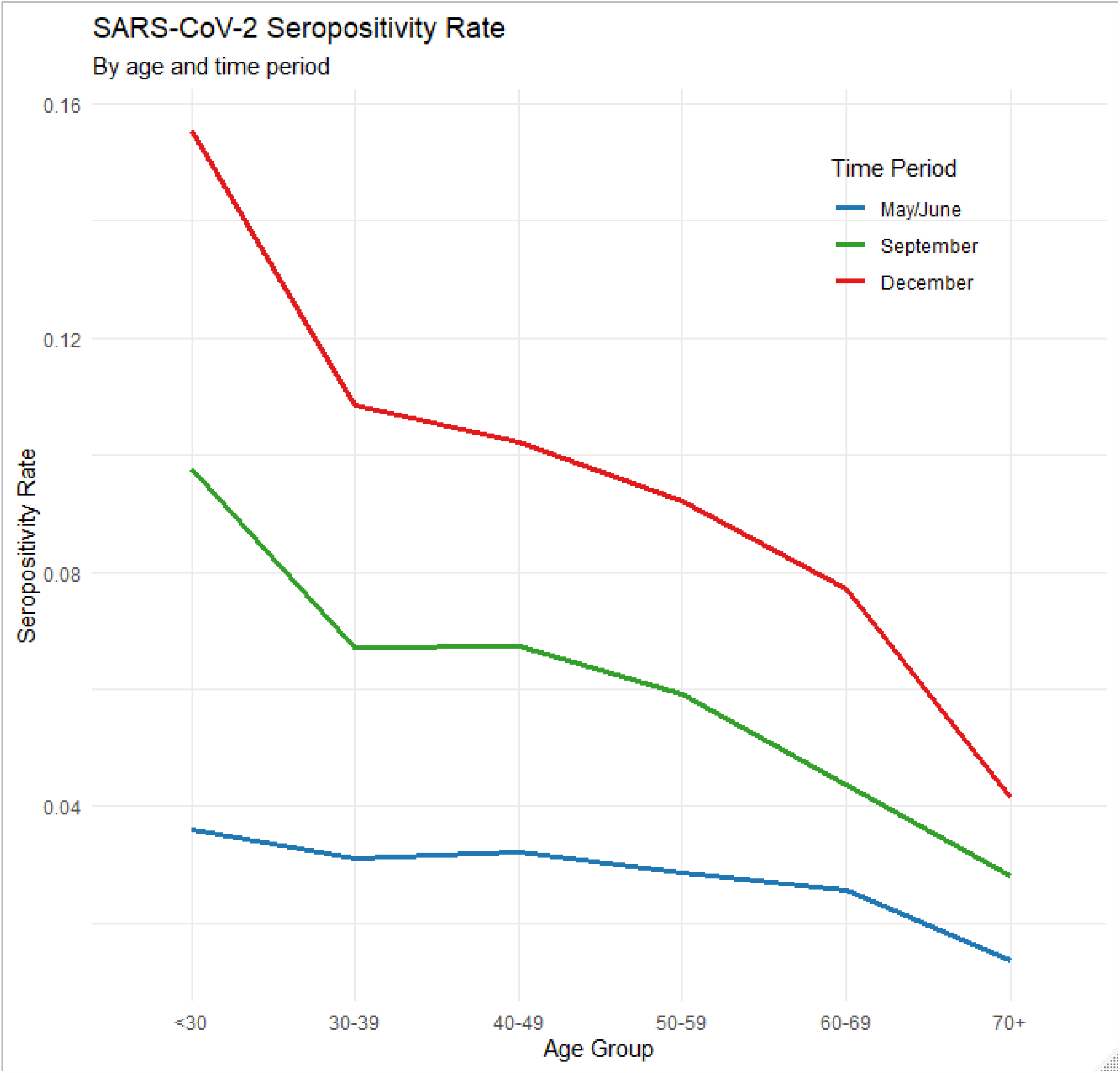

The overall time trend showed 3.0% seropositivity in May/June, 6.6% in September and 10.4% in December. For the purpose of generalizing to the US population, only the December time frame was considered. The estimate total SARS-CoV-2 infections was 26.2 million with a 95% CI based on 5000 bootstrap resamples of 24.9-27.5 million, or nearly twice the number of cases reported to the CDC as of December 6^th^.

## Discussion

This study estimated the seroprevalence of SARS-CoV-2 antibodies in a geographically diverse sample of adults over 3 distinct time periods in the latter half of 2020. The rate of positivity ranged from 1.4% to 7.9% depending on timeframe and age group. Our results suggest that many more infections have occurred than have been reported. This is likely due to asymptomatic or minimally symptomatic infections for which care was not sought or symptomatic infection for which testing was not obtained.

Various studies have been published, both before and after peer review, which have reported seroprevalence of SARS-CoV-2 antibodies in the US. Some evaluated only specific populations, such as those being admitted to hospital^6^, nursing home residents^7^, healthcare personnel, and hospital employees^8^. Other studies, including a previous prepublicaton from our group^9^, have evaluated samples from more general community populations. Bajema et al^10^, in collaboration with the CDC, evaluated over 177,000 samples from subjects in the community who were not being specifically evaluated for COVID-19 illness. They found an overall prevalence of under 10% in the most recent time period (September). Havers et al^11^ evaluated a convenience sample (n=16,025) of serological tests on sera submitted to 2 commercial laboratories from 10 US regions. Their estimates of seroprevalence ranged from 1% to 7%, with the highest rates occurring in the New York metro area, Louisiana and Connecticut. The timeframe of this testing differed by region and was earlier than the current study. The authors estimated that the seroprevalence implied that between 6 and 24 times the number of infections had occurred in the studied regions than had been reported.

Early studies that included age showed a higher prevalence in older patients^12^. Older age is associated with more severe disease with a higher probability of being identified as infected. We find the opposite trend, with an increasing prevalence in the younger population with an almost linear decrease in seroprevalence with increasing age. By far the lowest prevalence is in the elderly. This may be because elderly are the most at risk of complications from COVID-19 and have been avoiding high risk behaviors. CDC has a guidance paper on reducing the risk for infection including advice for people with health impairments^13^

Weaknesses of the study include the imbalanced representation of the US states, as well as the lack of samples from those under age 20 or over age 80. The age distribution is also more heavily weighted to the young adult years, which is not representative of the US population. Although the sample size was large, it was not large enough to stratify by both age and geography when estimating population seroprevalence. Finally, the life insurance-buying population tends to be both healthier and wealthier than average, and this could also bias the results in an indeterminate direction.

## Conclusion

The rate of SARS-CoV-2 seropositivity in this population of insurance applicants implies a burden of infection approximately twice the number of reported cases, with significant escalations over time, and a higher rate of prevalent infection among younger individuals.

## Data Availability

The data set is available to all not-for-profit releaser's on request.

## Notes

### Competing Interest Statement

The authors have declared no competing interest.

### Clinical Trial

Not a clinical trial

### Funding Statement

Self-funded research study

### Author Declarations

This study design was approved by the Western Institutional Review Board and determined to be exempt under the Common Rule and applicable guidance and determined it is exempt under 45 CFR 46.104(d)(4) using de-identified study samples for epidemiologic investigation, WIRB Work Order #1-1324846-1.

